# Factors Associated with Arteriovenous Fistula Maturation Failure among Patients Undergoing Hemodialysis in Hospitals Based in a Low and Middle-Income Country

**DOI:** 10.1101/2023.04.14.23288585

**Authors:** Arlon S Sichona, Victor Meza Kyaruzi, Alex Joseph, Maurice P Mavura, Ramadhani H Khamis

**Author notes:** **Correspondence to:** Dr Victor Meza Kyaruzi, MD, MMED, Department of Surgery, School of Medicine, Muhimhimbili University of Health and Allied Sciences, Dar es Salaam, Tanzania.

## Abstract

**Introduction:** The demand for haemodialysis among patients with end stage renal disease (ESRD) is rising worldwide, arteriovenous fistulas (AVF) are considered the gold standard vascular access modality for hemodialysis (HD) because of its longer patency, enhanced durability, and reduced risk of infection for those that mature compared to grafts and central venous catheters. This study will therefore assess the factors associated with arteriovenous fistula maturation for patients requiring hemodialysis in Dar es Salaam.

**Methods:** A multi-centre based prospective study conducted among patients with ESRD between April 2021 to May 2022 at Muhimbili National Hospital (MNH), Jakaya Kikwete Cardiac Institute (JKCI), Comprehensive Community Based Rehabilitation in Tanzania (CCBRT),Kairuki hospital (KH) in Dar es Salaam. Patients with End stage renal disease eligible for arteriovenous fistula (AVF) creation and Hemodialysis were included. Socio-demographic data were extracted from the patients, intraoperative and postoperative variables were obtained from medical records. Patients were assessed through eight weeks for maturation and complications. Data analyzed accordingly by IBM-SPSS version 27.0, Univariate and multivariate analysis were used to stratify the factors and control the confounders for the effects on outcome, and p-value of <5% was used to state the level of significance at 95% CI.

**Results:** Total of 151 fistulas were created, most (80.8%) were created on male, (31.8%) in the age group of less than 50 Years old, 58.9% of the participants had normal Body Mass Index (BMI), with majority (45%) being suffered from Hypertension and 54% of them were using Antihypertensive medication. Brachial cephalic fistulas were the most common type of fistula created 77(51%) and most matured 67(52.7%) among ESRD patients who had AVF creation, while radial cephalic fistulas were the most failed created AVFs 13 (54%). The AVF maturation failure rate was 16%.

**Conculusion:** Failure of newly created AVF is a major barrier to the successful establishment of hemodialysis access, in this study the failure rate of AVF maturation was 16 percent. The major factors associated with failure to mature were; extreme age group (50-59), being suffered with both hypertension and diabetes, long period of alcohol taking and distal location of AVF(radiocephalic fistula).Vascular imaging was not regularly done to assess the status of veins and arteries for AVF creation.

## INTRODUCTION

Chronic kidney disease (CKD) has been highlighted as a serious global public health concern. The number of end stage renal disease (ESRD) patients who require renal replacement therapy is expected to range between 4.902 and 7.083 million globally.(5) In Developed world it affects 10 -16% of the adult population. (6) Data from USA in 2016 indicate only 700,000 patients with ESRD require Hemodialysis on waiting list for renal transplant however, only 14% receive renal transplantation annually. (7) Population study report from India has noted that the Prevalence of CKD was 17.2% with country incidence of ESRD to be 229 per million population and >100,000 new cases start renal replacement therapy yearly. (8) In Sub-Saharan Africa the incidence of ESRD varies e.g. the incidence of ESRD was found to be 49.9 per million population in free state South Africa however, it was estimated that the incidence of ESRD accounted 13.9 per million population in Uganda.(9) (10) In Tanzania it was reported that the incidence of ESRD was 12.4 per million population, however the burden of those who requires hemodialysis annually have been increasing with few patients who benefits from renal transplant services.(11)

Due to poor transplant services in our region most of patients requires maintenance hemodialysis (HD) which is achieved with well-functioning vascular access (VA). An arteriovenous fistula (AVF), an arteriovenous graft (AVG), or central venous catheters (CVC) are all examples of vascular access (12). Arterio-venous fistula is simple, cost effective, and permanent mode of attaining hemodialysis among patient with ESRD. It was strongly recommended by studies that arteriovenous fistula as a crucial means of maintenance hemodialysis since it is related to a decreased mortality and helps patients to avoid catheter-related problems.(12) Therefore current practices promote use of AVF among hemodialysis patients. (4) Given the low risk of complications and great long-term patency rates, current clinical practice recommendations advocate using an AVF as the recommended vascular access for hemodialysis. Unfortunately, establishing and maintaining an AVF remains difficult and a significant source of morbidity in patients requiring HD.(13)

Despite of greater benefit of AVF, the procedure is not without adverse outcomes. Functional maturation failure was considered to be 35 per 100 patients who undergo AVF-creation at 8 weeks and resultant complications were aneurysm(11.1%), bleeding(22.2%), vascular thrombosis(5.6%) and stenosis(27.8%), surgical site infection(11.1%) and edema(22.2%) as described by Mtaturu et al 2020 (14). Also, Chan and others 2018 noted a maturation failure of 38 per 100patients at three months after AVF creation however, age, fistula location and vein size was independent factor for failure of maturation. However, AVF creation was been commonly practiced among patients with ESRD in our community the adverse outcomes associated with this operation and the factors which influence high index maturation failure at our centres had been not evaluated.

This study was therefore proposed to be performed to determine arteriovenous fistula maturation rate and factors associated among patients undergoing hemodialysis in Dar es Salaam, in order to create awareness and understanding on the factors associated with arterio-venous fistula maturation, and provide relevant information to health care providers and other important stakeholders on how to treat CKD patients and develop solutions.

## Methodology

A multicenter prospective study was conducted to look the relationship between exposures and outcomes at the same time. This study employed quantitative research methods in assessing the factors associated with arteriovenous fistula maturation in patients requiring hemodialysis in Dar es salaam, Tanzania. Study was carried out in the selected centers that are offering an AVF creation services includes Muhimbili National Hospital, JKCI, CCBRT and Kairuki hospital. These centres were selected because of high flow of patients with ESRD, well equiped facilities with hemodialysis machines, standard theatres and well trained surgeons for AVF creation services. Sample size was determined from study done 2020 at Muhimbili National hospital by Mtaturu et al where by a prevalence of functional maturation rate of 64% at a study power of >80% P 5%, 95%CI and a sample size of 151 patients was estimsted to be standard for the study. The source of data for this study was blood marker (creatine, urea), location of the fistula and the presence of central venous catheters and primary data was collected from all Patients with End stage renal disease seeking for AVF creation, identify Social demographic factors and Patient/clinical related factors associated with maturation of arterio-venous fistula for hemodialysis, maturation was assessed physically after 6 weeks, doppler ultrasound was not routine done to assess maturation, ability to sustain a two needle cannulation with flow rate of 200 ml/min succesifully was termed as functional Maturation. The electronic data were used to ascertain if patient had vascular image done before or after fistula creation and also confirm some of the complications that occurred after surgery. The collected data were processed, cleaned, coded, and entered before being imported into IBM SPSS version 24.0 for analysis. Percentages, tables, and graphs were used to summarize the data. To investigate the parameters related with arteriovenous fistula maturation, logistic regression analysis was used to calculate crude and adjusted odds ratios with 95percent confidence intervals (CIs). In the multivariate logistic regression model, variables with a p-value less than 0.05 were considered statistically significant. Adjusted Odds ratios with a 95% confidence interval were used for the interpretation of the factors associated with arteriovenous fistula maturation in patients requiring hemodialysis in Dar es salaam, Tanzania

## Results

Between April 2021 to May 2022, a total of 151 patients with ESRD underwent AVF creation at the fore mentioned institutions. Majority were male with male to female (M:F) ratio of 4.2:1. The age range was (20-84) years, with a mean age of 54.1± 14.2 years. Most of the participant was aged below 50 years at time of data collection. 67(44.4%) were from the technical or professional individual group and 89 (58.9%) of patients had normal BMI **table 1**. The prevalence of AVF maturation failure is 16 % as shown in **figure 1**.

**1:** Sociodemographic and patients characteristics.

1: Sociodemographic and patients characteristics
| Demographic Variables | Frequency | Matured | Not Matured | P- Value |
| --- | --- | --- | --- | --- |
| **Sex** |  |  |  |  |
| Male | 122(80.8) | 102(83.6) | 20(16.4) | 0.731 |
| Female | 29(19.2) | 25(86.2) | 4(13.8) |  |
| Total | 151 | 127(84.1) | 24(15.9) |  |
| **Age group** |  |  |  |  |
| <50 | 48(31.8) | 39(81.2) | 9(18.2) | 0.038 |
| 50-59 | 43(28.5) | 34(79.1) | 9(20.9) |  |
| 60-69 | 44(29.1) | 40(90.9) | 4(9.1) |  |
| 70+ | 16(10.6) | 14(84.1) | 24(15.9) |  |
| **BMI** |  |  |  |  |
| Underweight | 3(2.0) | 2(66.7) | 1(33.3) | 0.295 |
| Normal | 89(58.9) | 76(85.4) | 13(14.6) |  |
| Overweight | 40(26.5) | 31(77.5) | 9(22.5) |  |
| Obese | 19(12.6) | 18(94.7) | 1(5.3) |  |
| **Smoking status** |  |  |  |  |
| Yes | 40 (26.5) | 34(85.0) | 6(15.0) | 0.857 |
| No | 111 (73.5) | 93(83.8) | 18(16.2) |  |
| **Alcohol intake status** |  |  |  |  |
| Yes | 85(56.3) | 70(82.4) | 15(17.6) | 0.049 |
| No | 66(43.7) | 57(86.4) | 9(13.6) |  |
| **Alcohol taking duration** |  |  |  |  |
| Unknown | 48(37.5) | 44(91.7) | 4(8.3) | 0.029 |
| 1-4 Years | 28(21.9) | 19(67.9) | 9(32.1) |  |
| 5-10 Years | 28(21.9) | 24(85.7) | 4(8.3) |  |
| **Occupation** |  |  |  |  |
| None | 30(19.9) | 26(86.7) | 4(13.3) | 0.908 |
| Technical/Professional | 67(44.4) | 57(85.1) | 10(14.9) |  |
| Farmers | 25(16.6) | 20(80) | 5(20) |  |
| Business | 29(19.2) | 24(82.7) | 5(17.3) |  |

**Figure 1:**
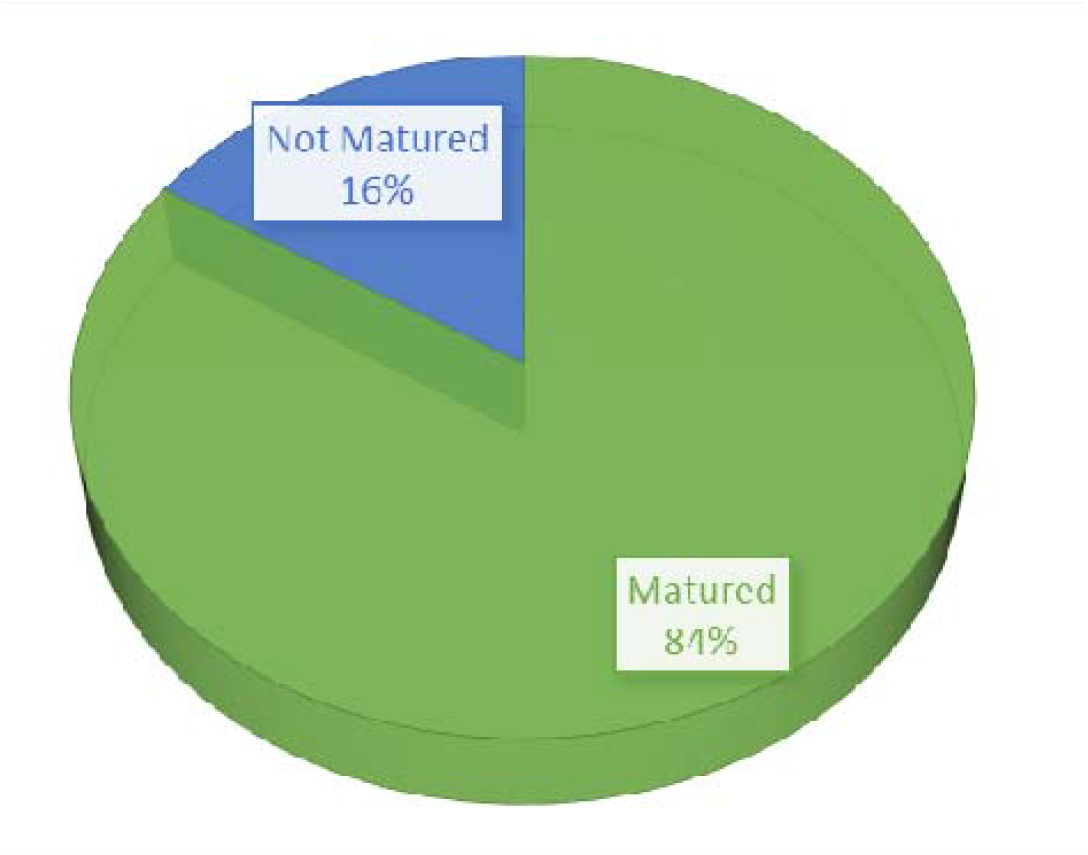
The Percentage of AVF maturation status among ESRD patients. The findings from figure 2 below, shows that, most participants 65.6% recorded non post AVF creation complications, swelling (9.9%), thrombosis (7.3%), stenosis (6.6%), bleeding (5.3%), limb edema (4.0%) and infection(1.3%)

**Figure 2:**
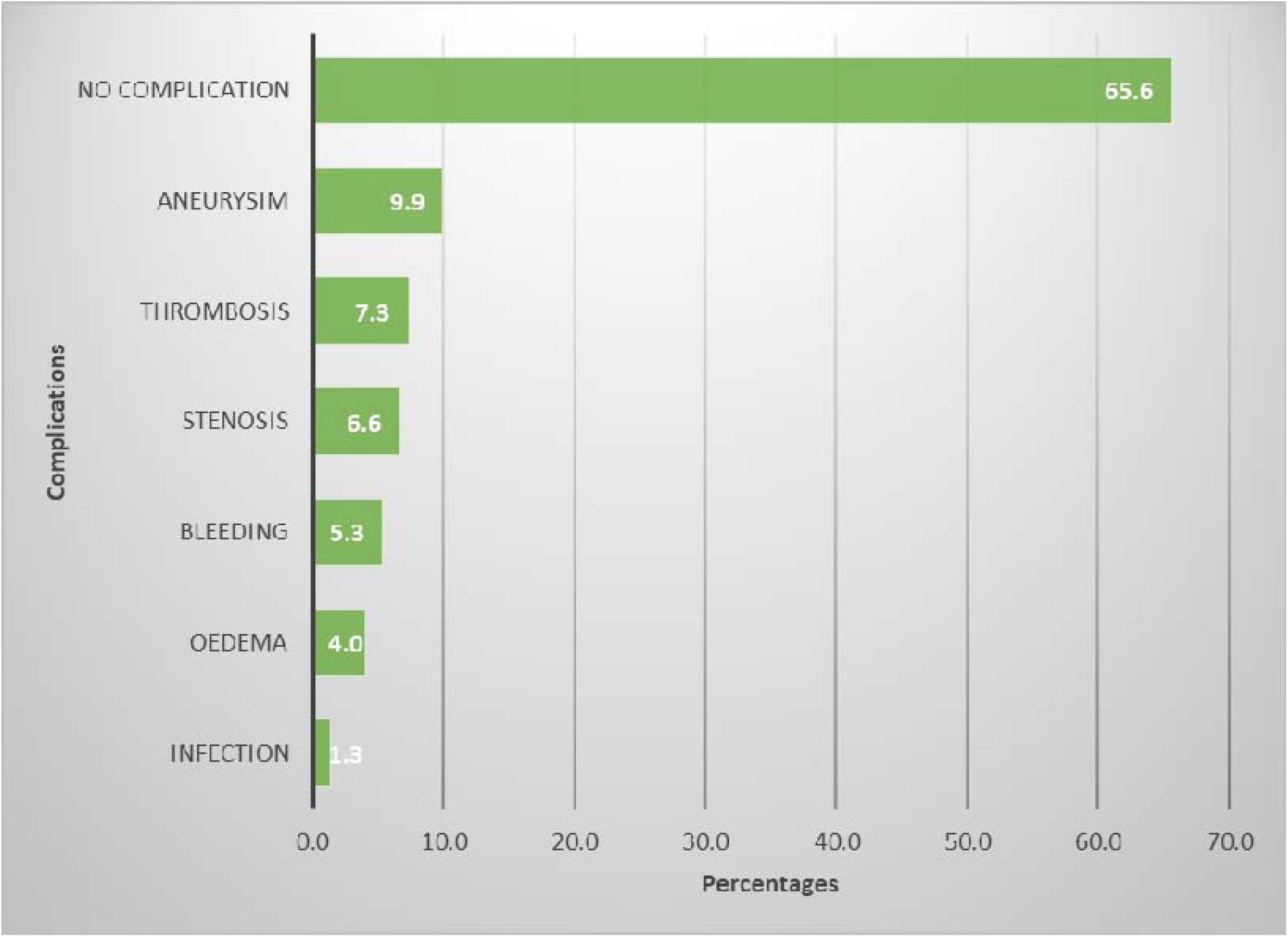
Complications post AVF creation.

### Baseline Clinical Characteristics

Majority, 16(15.8%) were diabetic and 68(45.0%) had hypertensive, however, 55(36.4%) had both hypertensive and diabetic cormobidities **table 1**. It was also, observed that 40(26.5%) of patients were smokers and 85 (56.3%) alcohol consumers. Lesser patients 29(19.6%) had elevated BUN and 90(59.3%) had high Creatinine level. Most of the fistula 77(51.0) were created on the Brachial-cephalic vascular access followed by radial-cephalic vascular access 57(37.7%) **table 2**.

**Table 2:**
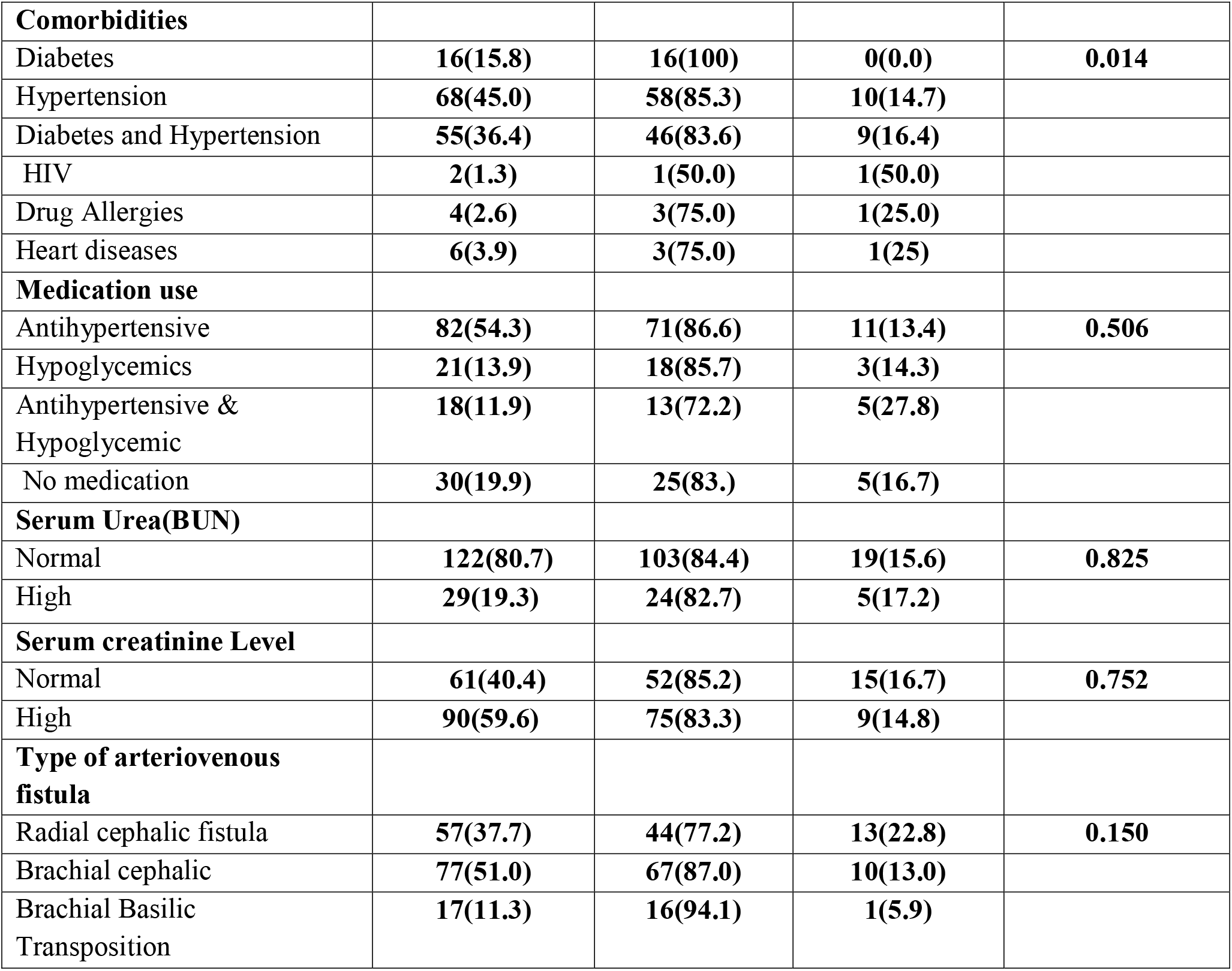
Baseline Clinical Characteristics.

### Risk factors for AVF maturation failure

Multiple factors including age, cormobidities, type of fistula creation, alcohol consumption duration and BMI were analysed for logistic multivariate analysis to control the confounders. The findings revealed that patients of age between 50-59, diabetes and hypertension cormobidity, having underwent radiocephalic fistula and those with history of alcohol consumption for more than 10 years had higher relative risk for AVF maturation failure with statistically significance adjusted odds ratio, **table 3**.

**Table 3:** Factors Associated with arteriovenous fistula maturation failure.

| Variable | COR (95% CI) | P - value | AOR (95% CI) | P – value |
| --- | --- | --- | --- | --- |
| **Age group** |  |  |  |  |
| <50 | Ref |  | Ref |  |
| 50-59 | 0.39 (0.32 – 0.60) | **<0.001** | 0.26 (0.96 – 0.77) | **0.038** |
| 60-69 | 2.32 (1.02 – 5.31) | 0.071 | 2.35 (0.85 – 6.49) | 0.093 |
| 70+ | 4.29 (1.88 – 9.83) | 0.093 | 3.53 (1.22 - 10.19) | 0.062 |
| **Comorbidities** |  |  |  |  |
| Diabetes | Ref |  | Ref |  |
| Hypertension | 1.16 (0.68 – 1.97) | 0.581 | 0.87 (0.48 – 1.58) | 0.652 |
| Diabetes and Hypertension | 0.28 (0.14 – 0.57) | 0.012 | 0.35 (0.16 – 0.77) | **0.014** |
| HIV | 0.75 (0.34 – 1.66) | 0.483 | 2.18 (0.76 – 6.28) | 0.152 |
| **Types of fistula** |  |  |  |  |
| Brachial cephalic | Ref |  | Ref |  |
| Radial cephalic | 0.39 (0.32 – 0.60) | 0.041 | 0.26 (0.06 – 0.77) | **0.049** |
| **Alcohol taking duration** |  |  |  |  |
| Unknown | Ref |  | Ref |  |
| 1-4 Years | 0.64 (0.42 – 0.97) | 0.047 | 0.82 (0.49 – 1.35) | 0.447 |
| 5-10 Years | 1.67 (0.99 – 2.80) | 0.063 | 1.41 (0.79 – 2.51) | 0.243 |
| More than 10 Years | 0.11 (0.10 – 0.32) | 0.041 | 0.01 (0.03 – 0.23) | **0.029** |
| **BMI** |  |  |  |  |
| Normal | Ref |  |  |  |
| Abnormal | 1.26 (0.52 – 3.03) | 0.605 | 1.40 (0.54 – 3.64) | 0.486 |

## Discusssion

The proper functioning of an AVF is critical for dialysis therapy, particularly with late stage renal disease. Patients with ESRD, an autologous fistula is the best VA option, with the lowest death rate and re intervention rate when compared to AVG and CVC, because of these benefits (33), the National Kidney Foundation of America recommended the use of native AVF, which has a primary success rate 65 percent amongst patients with ESRD who are ready to start HD(34). As more patients with ESRD are identified, the requirement to secure access becomes increasingly critical (35).

### The proportions AVF maturation among ESRD patients

According to this study, 16 percent of AVF created failed to mature for hemodialysis access. The percentage of patients with failed maturation were lower than the 20 percent to 54 percent of individuals with primary AVF failure described in a research conducted in African by Ashley Irish et al (21).

The percentages of failed AVF are crucial for identifying ESRD patients, understanding the variables related with AVF maturation, and provide valuable information to health care providers and other key stakeholders in the treatment of individuals with end-stage renal disease

The differences in the proportions of patients with failed AVF maturation between this study and other studies could be due to significant differences in blood flow rates between countries, which could explain the lower threshold for arteriovenous fistula failure in those countries with higher blood flow rates, significant differences in the dialysis population’s comorbidities, access to pre-dialysis CKD care, and the organization of end stage renal disease care among these ESRD patients (36).

### Age

According to this study patients with higher age group from 50-59 years old were likely to have failed maturation. The higher age group of 50 -59 years old being linked with failed maturation might be because of the age range that is more likely to have hypertension, diabetes mellitus (DM), and peripheral vascular disorders (PVD), the predominant age group of 50-59 years discovered in this study falls within the prominent age range of 41 to 60 years reported in a prior research done in Tanzania (37-38). This study stated main age group corresponds to the age group indicated by Pisoni et al in research carried out in Europe and the United States (39). The reasons for the similarities in the prevalent age group between this study and other European studies point to a shift in the epidemiological pattern of illnesses in Sub-Saharan Africa as a result of an increase in NCDs, potentially related to westernization of lifestyles (40).

These findings are also corroborated by prior findings from a European research, which found that age is a decisive factor in AVF maturation, with older patients (e.g., over 65 years) having lower patency rates.(2). Former research has linked older age groups to longer maturation rate and lower rates of effective AVF use, with patients aged 44 having the shortest maturation times and the highest percentage of successful AVF use. (3). Furthermore, the findings of this study are similar to those of a previous study, which revealed that AVF maturation delays and greater failure rates were all associated to age, the need for assistance, and institutionalization, Patricia Barreto et al (3).

AVF patency in old and younger persons has been studied, and the results suggest that elderly people with radiocephalic AVF have a higher main failure rate and lesser patency (43-46). This contradicts the findings of Lok et al, who suggested that age should not be a limiting factor when evaluating vascular access choices for HD because their study indicated that patients over and under 65 years of age had identical survival and procedure rates. (3).

In our study the mean age of patients with ESRD was 54.1 years, with a range of 20 to 84 years. This research’s findings contrast those of a previous study conducted in Tanzania, which showed a mean age of 47 years and a range of 18 to 69 years (22). The variations in mean age and age range between this study and the prior study might be attributable to changes in the nature of the study’s population.

### Hypertension and diabetes

Patients with hypertension and diabetes were more likely to have AVF failure rate than those with other Noncommunicable Diseases, according to the research. The findings of this study reflect those of a previous study, which found that the number of obese patients with ESRD who also have type 2 diabetes is steadily increasing and is significantly linked to a higher rate of AVF failure because they are predisposed to arteriosclerosis and vessels in the forearm are more difficult to reach due to thick fat tissue.(47-48). Other studies, on the other hand, found that hypertension was not a significant risk factor for AVF maturation. Kim et al. recently conducted a single-center cohort study (38 men and 12 women) to evaluate patient characteristics, vein diameter, and distensibility of the radiocephalic vein at the wrist. There was no evidence of a link between AVF maturation and patient variables such as hypertension, diabetes, or gender in the study. (4).

### Alcohol taking and Duration of alcohol taking

Patients who used alcohol for a longer period of time were shown to have a higher risk of AVF failure rate than those who did not consume alcohol. The findings of this study corroborate those of a prior study, which found that alcohol takers on hemodialysis had a higher rate of late AVF failure. The differences in the results might be due to the fact that previous peripheral vascular damage in past and present alcohol takers can produce acute AVF blockage (49).

### Types of Fistulas (AVF)

When compared to other forms of fistulas, radial cephalic fistula was the most prevalent among patients with AVF failures, according to the research. This study’s findings are consistent with prior research, which found that older patients with radiocephalic AVF had a greater main failure rate and worse patency (50). This conclusion might be due to the fact that the location of an AVF increases the chance of cannulation failure, especially in individuals with radiocephalic AVFs (51) However, the complexity of the processes of effectively establishing AVF formation among patients limits these findings, since several practices-related aspects such as surgical and cannulation technique and competence are difficult to standardize assess accuracy and consistency across different settings (52). Failure of the AVF to mature is a major cause of death and morbidity in people on dialysis. Other factors such as smoking, blood markers (creatinine and Urea) were not statistically significant in this study.

### Strength and Limitation of the Study

Well-characterization of the study population and ensuring standardized and accurate collection of patient and surgical variables were the strength of this study.

The limitation of this study were the limited number of participants and events, insufficient granularity of data (such as severity of comorbidities, location of the CVC in relation to the AVF and lack of standardized blood pressure.

The other limitation is that, AVFs placed prior to initiation of dialysis are not directly addressed by this study and merit further investigation.

## Conclusion and recommendation

### Conclusion

ESRD were more observed in male, aged below 50 years, normal BMI, hypertensive, and college/university level education. In this study the failure rate among ESRD patients requiring hemodialysis were 16 percent. Advanced age, being suffered with hypertensive and diabetic, alcoholism for more than 10 years were significant independent factors for AVF maturation failure. Radiocephalic created fistula was significantly associated with high failure rate in this population cohort Failure of newly created AVF is a major barrier to the successful establishment of hemodialysis access.

### Recommendation

1. Pre-operative ultrasonographic scan (US) mapping of the Arteries and Veins will help to demonstrates the association of vessel diameter and distensibility as well as AVF maturation and patency rates.
2. Further studies are needed to determine the cause of AVF maturation failure among elderly, hypertensive and diabetics, and patients with chronic alcohol use.

## Data Availability

All data produced in the present study are available upon reasonable request to the authors

## DECLARATIONS

### Ethics approval and consent to participate

Ethical clearance for the study was obtained from Muhimbili University of Health and Allied Sciences (MUHAS) IRB REF DA.282/298/01.C. Separate permissions were sought from Muhimbili National Hospital (MNH), Jakaya Kikwete Cardiac Institute (JKCI), Comprehensive Community Based Rehabilitation in Tanzania (CCBRT) and Kairuki Hospital (KH) to conduct the study in the health facilities.

### Consent for publication

Verbal Informed consent for participation and publication of results was obtained from study participants following approval by MUHAS institution review board.

### Availability of data and materials

All data and materials shall be made available as shall be requested

### Authors’ Contributions

Conception and design Sichona A, Kyaruzi VM, Joseph A, Mavura MP, Khamis RH, All authors have approved the submitted manuscript.

